# Factors associated with unintended pregnancies among unmarried adolescents and the potential of using mobile money shops: A Cross-sectional study in Eastern Uganda

**DOI:** 10.1101/2025.01.17.25320704

**Authors:** Makiko Komasawa, Miho Sato, Kiyoko Saito, Sumihisa Honda, Robert Ssekitoleko, Peter Waiswa, Kharim Mwebaza Muluya, Sheba Gitta, Myo Nyein Aung

**Author notes:** **Institutional address:** Ogata Sadako Research Institute for Peace and Development, Japan International Cooperation Agency (JICA), 10-5, Ichigaya-honmuracho, Shinjuku-ku, 162-8433, Tokyo, Japan. **Corresponding author:** Makiko Komasawa, Ogata Sadako Research Institute for Peace and Development, Japan International Cooperation Agency (JICA), 10-5, Ichigaya-honmuracho, Shinjuku-ku, 162-8433, Tokyo, Japan,.

## Abstract

**Background:** Adolescent sexual and reproductive health and rights (SRHR) remains a critical public health issue in low- and middle-income countries, with adverse health, educational, and economic consequences. Uganda faces significant challenges with high rates of unintended pregnancies among adolescents. This study investigated the factors associated with unintended pregnancies among unmarried adolescents in Eastern Uganda and explored the potential of using mobile money vendors to provide SRHR services.

**Methods:** A cross-sectional study was conducted among 1267 unmarried girls/boys, aged 15-19 years, in the Busoga region. Data were collected through face-to-face interviews, using a structured questionnaire. Sociodemographic characteristics, sexual behaviors, contraceptive knowledge, and environmental factors were analyzed. Multivariate logistic regression analysis identified factors associated with unintended pregnancies.

**Results:** Of the adolescents who had sexual intercourse (n=876), 22.5% (n=197) experienced unintended pregnancies. Fundamental factors associated with lower prevalence of pregnancy included being a current student (adjusted odds ratio [AOR]: 0.36, 95% confidence interval [CI]: 0.24-0.54), living with parents (AOR: 0.48, 95% CI: 0.33-0.69), and teacher engagement (AOR: 0.56, 95% CI: 0.37-0.84). Factors associated with higher prevalence of pregnancy included knowledge regarding contraceptive use (AOR: 2.28, 95% CI: 1.16-4.49), parental communication (AOR: 1.91, 95% CI: 1.32-2.75), parental contraception support (AOR: 1.64, 95% CI: 1.03-2.59), and mobile phone possession (AOR: 2.31, 95% CI: 1.09-4.90). Public health facilities and community-based distributors, including mobile money vendors, were considered comfortable channels for obtaining contraceptives for adolescents who had experienced pregnancy.

**Conclusions:** Unintended pregnancies among unmarried adolescents in Eastern Uganda were influenced by educational, parental, and environmental factors. Enhancing SRHR education from earlier ages, parental involvement, and leveraging community resources such as mobile money vendors could improve adolescent SRHR outcomes. Innovative approaches outside conventional health and education sectors are necessary for sustainable and effective adolescent SRHR programs.

**Trial registration:** This study was registered at Japan’s University Hospital Medical Information Network (UMIN000053332) on 12 January 2024.

**Plain English summary:** Adolescent sexual and reproductive health and rights (SRHR) is a pressing public health issue globally. Uganda has a high rate of unintended pregnancies among adolescents. This study investigated the factors associated with unintended pregnancies among unmarried adolescents in Eastern Uganda and explored the potential of using mobile money vendors to provide SRHR services.

This cross-sectional study was conducted using a structured questionnaire at 60 vendors. It included 1267 unmarried girls/boys aged 15-19 years. Of 876 adolescents who ever had sexual intercourse, 22.5% experienced unintended pregnancies. Factors associated with a lower prevalence of pregnancy were: a current student (adjusted odds ratio [AOR] = 0.36), living with parents (AOR = 0.48), and teacher engagement (AOR = 0.56). Contrarily, factors associated with a higher prevalence of pregnancy were: knowledge regarding contraceptive use (AOR = 2.28), parental communication (AOR = 1.91), parental contraception support (AOR =1.64), and mobile phone possession (AOR = 2.30), which may imply that teenagers who had experienced pregnancy tend to become more serious about contraception and engagement of parents and teachers enhanced. In addition, community-based SRHR service distributors, including mobile money vendors, were considered comfortable channels.

In conclusion, unintended pregnancies among unmarried adolescents in Eastern Uganda were influenced by educational, parental, and environmental factors. Enhancing SRHR education before starting sexually active and parental and school involvement may reduce unintended pregnancies among unmarried adolescents in Eastern Uganda. In addition, leveraging accessible community-based distributors, such as mobile money vendors, may be a potential channel for delivering SRHR information and contraceptives.

## Background

Adolescents sexual and reproductive health and rights (SRHR) is a pressing public health issue globally. In low- and middle-income countries, approximately 21 million girls aged 15-19 years become pregnant and almost half of these pregnancies are unintended [1]. These early pregnancies not only contribute to maternal and child mortality but also lead to unsafe abortions [1, 2]. The impact of early pregnancy extends beyond health, affecting a girl’s education and income-earning potential and perpetuating a cycle of poverty that can last for generations [3, 4]. This situation is a result of unmet needs for SRHR services among young populations [5, 6].

Sub-Saharan Africa (SSA) has high rates of adolescent pregnancies, and this trend is expected to increase [7]. Among the 29 SSA countries, Uganda ranks the fifth with a high prevalence of unintended pregnancies [8]. Many girls engage in sexual activity before marriage. The median age for the first sexual relationship among women aged 25-49 years was 16.9 years in 2021 [9]. The maternal mortality rate remains high at 189 per 100,000 live births, with 17% of these involving girls aged 15-19 years [2, 9]. The rate of any contraceptive use among all women aged 15-49 year remains at 44.0% and for modern methods at 40.1% [9]. Ninety-seven percent of pregnant female students drop out of school, despite the education policy encouraging students who have given birth to return to school [10]. Additionally, there has been an increase in the number of unintended pregnancies resulting from "transactional sex," where girls economically affected by coronavirus disease 2019 crisis engage in sex in exchange for small amounts of financial aid or transportation [11]. There remains a need to improve the quality and quantity of public SRHR services in Uganda, including youth-friendly family planning services [12, 13]. The Ugandan government has implemented various policies and programs over the past 15 years, such as expanding the compulsory education period, enacting defilement laws, prohibiting marriage for girls under 18 years of age, prohibiting sexual intercourse with girls under 18 years of age, and promoting adolescent contraception [11, 14]. However, adolescent pregnancy rates have remained stagnant over the past decade [14].

Since the International Conference on Population and Development in 1994, numerous interventions have been undertaken to improve adolescent SRHR [15]. Most of these interventions have been conducted in schools, primary healthcare facilities, and communities by peer-educators [16-18]. However, some studies highlighted that many programs have not adequately reached unmarried teenagers who need SRHR services [16, 19]. Additionally, the number of SRHR services has been limited, and their adoption has been insufficient, making many of these programs unsustainable and ineffective despite substantial investments [19]. Consequently, youth SRHR programs have stagnated for almost two decades [6, 20]. In light of these challenges, several studies have stressed the need for innovative approaches to ensure sustainability and scalability [17, 18, 21, 22].

In our exploration of effective approaches in Uganda, we focused on utilizing mobile money shops (vendors) as a channel to connect with adolescents and provide them with SRHR services [23]. These vendors are available throughout the country; as in 2023, 63% of Uganda’s population was using mobile phones [24]. The number of registered mobile phone customers increased by 1.3 times from 2015 to 2019 [25]. Our preliminary study also discovered that 85% of adolescents accessed vendors more than weekly to use various services such as money transfers, purchasing airtime/data, and making cash deposits [23].

Given the situation in Uganda, we initiated a hybrid effectiveness-implementation trial to reach teenagers using mobile money shops to reduce unintended pregnancies [23]. Based on baseline data collected as part of the trial, this study aimed to explore the current situation of sexual behavior and determine factors associated with unintended pregnancy among adolescents aged 15-19 years. More importantly, this study assessed the potential of an innovative approach based on vendors in Uganda.

## Methods

### Study design and setting

The detailed study design has been published elsewhere [23] and was registered at J アパ根セ ‘the University Hospital Medical Information Network (UMIN000053332) in Japan. Here, we provide the fundamental parts of the study design for interpreting the study results. This was a cross-sectional study of unmarried adolescents aged 15-19 years in the Busoga region of Eastern Uganda. The center of the Busoga region is approximately 80 km from Kampala, the capital city. Two municipalities in the Busoga region with similar characteristics were selected: Iganga and Bugiri. In 2022, the population of Iganga municipality was 55,263 and that of Bugiri was 28,747, whose teenage population (10-17 years old) accounted for 20.9% and 19.3%, respectively. We selected 30 vendors from each municipality according to predefined study criteria. Owners of mobile money businesses (vendors) aged 18-30 years, who have operated for more than one year, and are willing to participate in the study were selected. Stand-alone shops were selected to secure the privacy of customers and commodities. The geographical distribution, diversity, population density, availabilities of health facilities and schools of each municipality were also considered. We recruited teenage customers from selected vendor sites. Recruitment ceased after reaching the desired sample size of approximately 20 per vendor.

### Participants

The eligible study participants were unmarried girls/boys aged 15-19 years who used the selected vendors in the target municipalities and had no plans to move out of the municipalities during the study period. Adolescents aged 13-14 years were excluded because they tended to be less sexually active and used fewer vendors.

We applied a non-randomized prospective controlled design because vendor customers cannot be assigned to specific vendors, and they choose several vendors at their own convenience. We decided on a sample size of 1200 for the intervention study in the two municipalities using a formula based on the available data and previous studies, using a calculation method explained elsewhere [23]. Adolescents under 18 years of age who agreed to contact their parents/guardians during the screening process were recruited.

### Data collection

A structured questionnaire was developed using research instruments based mainly on the 2016 Uganda Demographic and Health Survey [26] and other studies conducted in SSA [27-33]. We have added questions regarding social and physical opportunities, and those related to vendors’ usage [23]. We developed a questionnaire in English and translated it into the local language. Before finalizing the questionnaire, we conducted a series of pre-tests with 26 adolescents (10 boys and 16 girls) and modified them for easy answering. Although most adolescents used English daily, some participants were comfortable using their local language to discuss sensitive topics in SRHR. We employed data collectors who could manage the local language and had academic backgrounds in the social welfare or health fields. The questionnaire included five categories: (1) sociodemographic characteristics, (2) sexual behavior, (3) contraception knowledge and capability, (4) environmental factors preventing pregnancy, and (5) preference for contraceptive practices.

The questionnaire was administered face-to-face by trained data collectors using preprogrammed tablet computers [23]. Interviews were conducted in a private location near vendors’ or teenagers’ homes between February and March 2024. Data collection lasted between 30 and 40 min per interview.

### Data analysis

The primary outcome was the experience of pregnancy experience of girls or impregnation of boys, regardless of sex. First, we described the sociodemographic characteristics of the participants and their sexual behaviors with the comparison of the two groups (pregnancy/impregnation [pregnant] versus never-pregnant) using the chi-square test. Second, we described the variables in each category using two-group comparisons. Third, multivariate logistic regression analysis was performed to identify the factors associated with the primary outcome. Variables that showed a statistically significant difference in bilateral regression were considered for multivariate logistic regression. In addition, we removed correlated variables and added the basic variables identified in a previous study, which accounted for 21 variables. Statistical significance was set at p < 0.05. Data analysis was conducted using STATA v.18.

### Ethical consideration

Ethical approval was obtained from the Ethics Review Committee of Uganda Christian University (UCUREC-2023-710), the Uganda National Council for Science and Technology (SS2284ES), and the Ethics Review Committee of the JICA Ogata Sadako Research Institute for Peace and Development in Japan (JICA Ogata RI, reference: JICA [DI] 202312220002). Written informed consent was obtained from all adolescent participants and the parents/guardians of adolescents aged 15-17 years. Adolescents may feel uncomfortable participating in the study as the content includes their sexual behaviors. To mitigate their anxiety, we informed them that their participation was truly voluntary without any harm and that they could withdraw their participation at any time, even after the survey and collected information were strictly confidentially managed. To obtain consent from parents/guardians of adolescents under 18 years of age, highly experienced and skilled researchers visited the homes of expected participants and explained the study to the parents/guardians.

## Results

We recruited 1281 eligible teenagers for the intervention [Additional file_01]. We excluded 14 participants who did not responded their pregnancy experiences, and 1267 participants were included in the study. Table 1 shows the sociodemographic characteristics of the participants in two groups: pregnant versus never-pregnant. Out of the analyzed participants, 69.1% had sexual intercourse (876/1267). Among the respondents who had sexual intercourse, 22.5% (197/876) had gotten pregnant. Of the respondents, boys were 52.5% and those aged below 18 years were 30.8%. More than half of them (57.7%) completed lower secondary school (senior 4). The highest proportion of religion was Islam (54.3%). We recruited both in- and out-of-school adolescents; 51.2% were currently in-school students and 39.8% were working. Only sex and religion were not statistically different between the two groups. Table 2 presents respondents’ sexual behaviors. Overall, 84.7% (1073/1267) ever had a girlfriend(s)/boyfriend(s) (i.e., intimate partner[s]). Among them, 81.6% (876/1073) had ever experienced sexual intercourse. The mean age at first intercourse was 15.39 years (standard deviation [SD]: 1.98) and was significantly different between the pregnant (mean: 15.04 years, SD: 2.01) and never-pregnant (mean: 15.48 years, SD: 1.96) groups (difference: 0.44 years, p < 0.0053). In the last four months, 88.7% (777/876) had intimate partners, and 84.7% (660/777) had sexual intercourse. Out of these, 81.5% (538/660) had taken contraceptives during intercourse in the last four months. Out of those who got pregnant, 51.0% (97/190, 7 did not responded about outcome of pregnancy, out of 197) had a live birth(s).

**Table 1.** Basic characteristics of participants(n = 1267)

| Variables | Total<br>(n=1267) |  | Pregnant<br>(n=197) |  | Never pregnant<br>(n=1070) |  | p-value |
| --- | --- | --- | --- | --- | --- | --- | --- |
|  | n | % | n | % | n | % |  |
| **1) Gender** |  |  |  |  |  |  | 0.321 |
| Boys | 665 | 52.5 | 97 | 49.2 | 568 | 53.1 |  |
| Girls | 602 | 47.5 | 100 | 50.8 | 502 | 46.9 |  |
| **2) Age** | | | | | | | $p < 0.001$ |
| 15-17 years old | 390 | 30.8 | 24 | 12.2 | 366 | 34.2 |  |
| 18-19 | 887 | 69.2 | 173 | 87.8 | 704 | 65.8 |  |
| **3) Education level completed** |  |  |  |  |  |  | 0.017 |
| Primary school or less | 429 | 33.9 | 83 | 42.1 | 346 | 32.3 |  |
| Lower secondary school | 731 | 57.7 | 103 | 52.3 | 628 | 58.7 |  |
| Secondary school or higher | 107 | 8.5 | 11 | 5.6 | 96 | 9.0 |  |
| **4) Religion** |  |  |  |  |  |  | 0.204 |
| Christianity | 327 | 25.8 | 54 | 27.4 | 273 | 25.5 |  |
| Islam | 688 | 54.3 | 113 | 57.4 | 575 | 53.7 |  |
| Others | 252 | 19.9 | 30 | 15.2 | 222 | 20.8 |  |
| **5) Currently in education** |  |  |  |  |  |  | p |
| Yes | 649 | 51.2 | 45 | 22.8 | 604 | 56.5 | p < 0.001 |
| No | 618 | 48.8 | 152 | 77.2 | 466 | 43.6 |  |
| **6) Currently in working** |  |  |  |  |  |  |  |
| Yes | 504 | 39.8 | 114 | 57.9 | 390 | 36.5 | p < 0.001 |
| No | 763 | 60.2 | 83 | 42.1 | 680 | 63.6 |  |

**Table 2.** Sexual behavior.

| Variables | Total |  | Pregnant |  | Never pregnant |  | p-value |
| --- | --- | --- | --- | --- | --- | --- | --- |
|  | n | % | n | % | n | % |  |
| **1) Ever had girl/boyfriends (n=1267)** |  |  |  |  |  |  |  |
| Yes | 1073 | 84.7 | 197 | 100.0 | 876 | 81.9 | p < 0.001 |
| No | 194 | 15.3 | 0 | 0.0 | 194 | 18.1 |  |
| **2) Ever had sexual intercourse (n=1073)** |  |  |  |  |  |  |  |
| Yes | 876 | 81.6 | 197 | 100.0 | 679 | 77.5 | p < 0.001 |
| No | 190 | 17.7 | 0 | 0.0 | 190 | 21.9 |  |
| Declined to answer | 7 | 0.7 | 0 | 0.0 | 7 | 0.8 |  |
| **3) Age at the first sexual intercourse (n=866)** |  |  |  |  |  |  |  |
| Age (SD) | 15.39 (1.98) |  | 15.04 (2.01) |  | 15.48 (1.96) |  | 0.005 |
| **4) Having girl/boy friends in the last four months (n=876)** |  |  |  |  |  |  |  |
| Yes | 777 | 88.7 | 180 | 91.4 | 597 | 87.9 | 0.394 |
| No | 89 | 10.2 | 15 | 7.6 | 74 | 10.9 |  |
| Declined to answer | 10 | 1.1 | 2 | 1.0 | 8 | 1.2 |  |
| **5) Sexual intercourse in the last four months (n = 777)** |  |  |  |  |  |  |  |
| Yes | 660 | 84.9 | 163 | 90.6 | 497 | 83.3 | 0.051 |
| No | 107 | 13.8 | 15 | 8.3 | 92 | 15.4 |  |
| Declined to answer | 10 | 1.3 | 2 | 1.1 | 8 | 1.3 |  |
| **6) Use of contraceptives in the last four months (n = 660)** |  |  |  |  |  |  |  |
| Yes | 538 | 81.5 | 127 | 77.9 | 411 | 82.7 | 0.384 |
| No | 116 | 17.6 | 34 | 20.9 | 82 | 16.5 |  |
| Declined to answer | 6 | 0.9 | 2 | 1.2 | 4 | 0.8 |  |

Table 3 shows factors related to contraception knowledge and capability. A total of 96.1% had heard any contraceptives, while the knowledge regarding how to use any contraceptives was lower (79.9%). Knowledge regarding the menstrual cycle and timing of pregnancy was low (overall 9.1%). Friends were the most frequent source of contraceptive information, followed by school, television/radio, and public health facilities. Partner agreement on contraceptive use was 80.8%, and differences between the two groups. Knowledge regarding how to use contraceptives and sources of contraceptive information (public health facility, private facility, school, community-based distributor, and parent/relatives) were significantly different between the pregnant and never-pregnant groups. Table 4 describes the environmental factors preventing pregnancy. Nearly 70% of the respondents lived with parents, showing a higher percentage of never-pregnant than the pregnant group (p < 0.001). Communication with parents/guardians about intimate partners was low among all respondents (27.6%). Perception of support by healthcare workers, teachers, and the community was relatively high among all respondents (82.7%, 68.4%, and 69.1%, respectively), with statistically significant differences between the two groups, except for community support. Regarding mobile phone possession, 81.2% of teenagers had it, with the pregnant group having significantly higher ownership than the never-pregnant group (p < 0.001). Regarding vendor service usage, 64.2% used vendor services at least once a day. The most frequent use of service was airtime/data for all, while the pregnant group showed a higher percentage of withdraws than the never-pregnant group. Table 5 shows the results of preferences on contraception practice. Desire to acquire knowledge on contraception and develop intimate relationships were high in both the pregnant and never-pregnant groups (92.9% and 83.2%, respectively). The most preferred source of SRHR information was the public health facility, and others were varied. Regarding comfortable channels of obtaining contraceptives, public health facilities were the most preferred source), followed by pharmacies) and private health facilities. Statistically significant differences between the two groups were: desire to develop intimate relationship, preferred source of SRHR information, some of channel of obtaining contraceptives. Nearly 70% expected vendors to be suitable channels for receiving SRHR information and contraceptives before the trial intervention started.

**Table 3.**
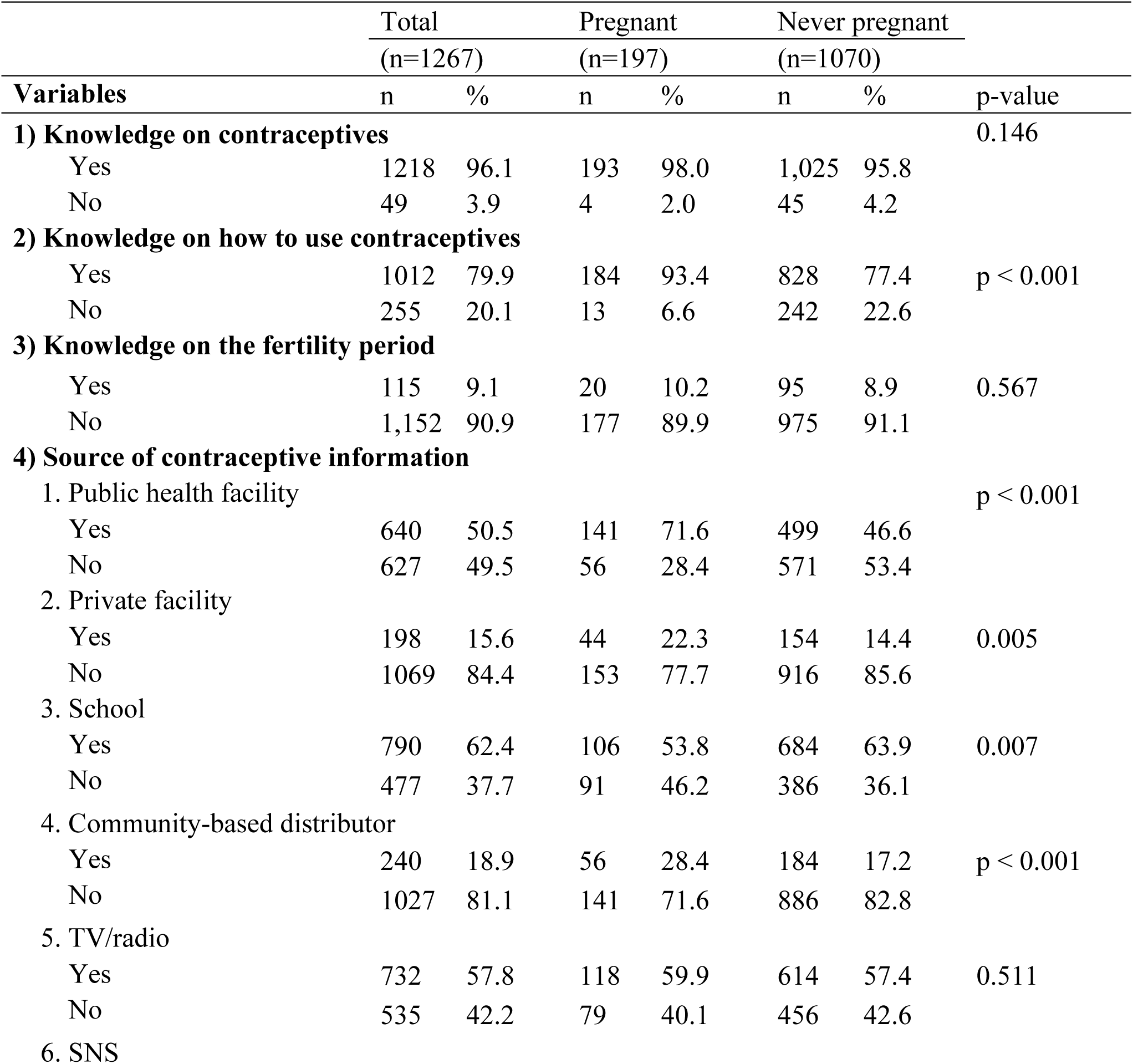
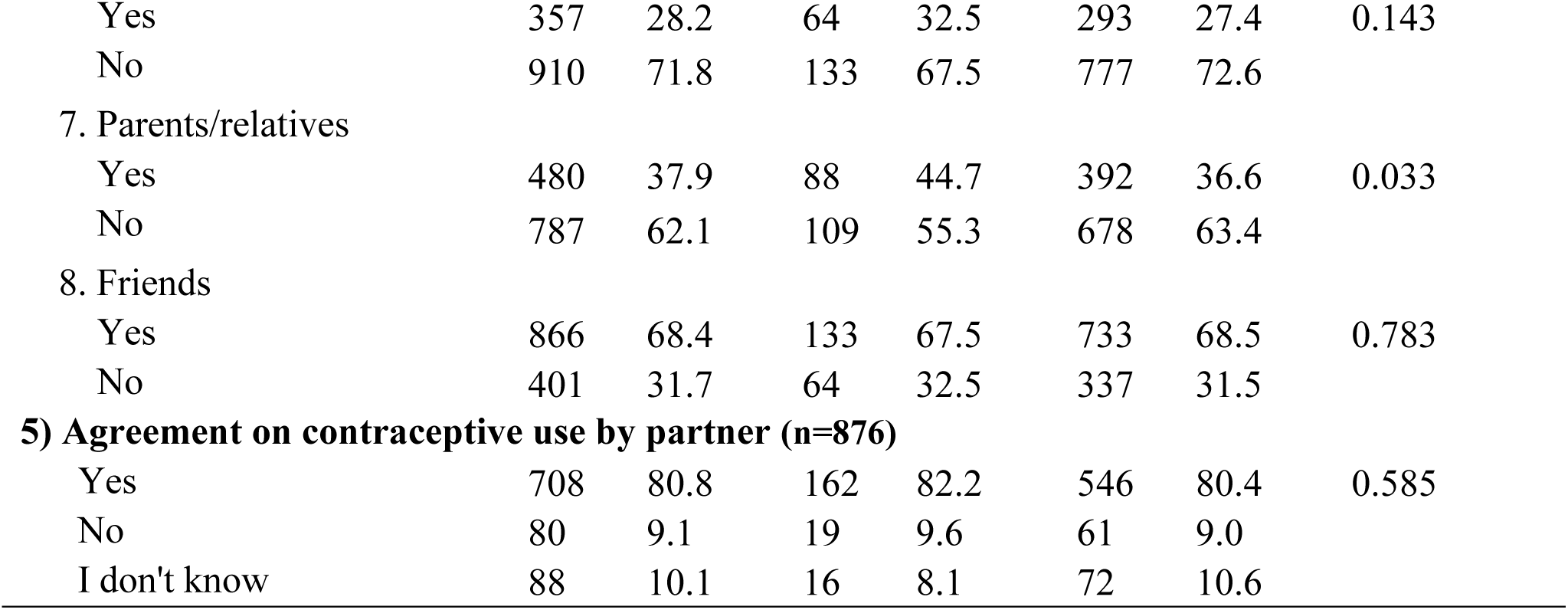
Knowledge and capability of contraception (n = 1267)

| Variables | Total<br>(n=1267) |  | Pregnant<br>(n=197) |  | Never pregnant<br>(n=1070) |  | p-value |
| --- | --- | --- | --- | --- | --- | --- | --- |
|  | n | % | n | % | n | % |  |
| **1) Knowledge on contraceptives** |  |  |  |  |  |  | 0.146 |
| Yes | 1218 | 96.1 | 193 | 98.0 | 1,025 | 95.8 |  |
| No | 49 | 3.9 | 4 | 2.0 | 45 | 4.2 |  |
| **2) Knowledge on how to use contraceptives** |  |  |  |  |  |  | p < 0.001 |
| Yes | 1012 | 79.9 | 184 | 93.4 | 828 | 77.4 |  |
| No | 255 | 20.1 | 13 | 6.6 | 242 | 22.6 |  |
| **3) Knowledge on the fertility period** |  |  |  |  |  |  | 0.567 |
| Yes | 115 | 9.1 | 20 | 10.2 | 95 | 8.9 |  |
| No | 1,152 | 90.9 | 177 | 89.9 | 975 | 91.1 |  |
| **4) Source of contraceptive information** |  |  |  |  |  |  |  |
| 1. Public health facility |  |  |  |  |  |  | p < 0.001 |
| Yes | 640 | 50.5 | 141 | 71.6 | 499 | 46.6 |  |
| No | 627 | 49.5 | 56 | 28.4 | 571 | 53.4 |  |
| 2. Private facility |  |  |  |  |  |  | 0.005 |
| Yes | 198 | 15.6 | 44 | 22.3 | 154 | 14.4 |  |
| No | 1069 | 84.4 | 153 | 77.7 | 916 | 85.6 |  |
| 3. School |  |  |  |  |  |  | 0.007 |
| Yes | 790 | 62.4 | 106 | 53.8 | 684 | 63.9 |  |
| No | 477 | 37.7 | 91 | 46.2 | 386 | 36.1 |  |
| 4. Community-based distributor |  |  |  |  |  |  | p < 0.001 |
| Yes | 240 | 18.9 | 56 | 28.4 | 184 | 17.2 |  |
| No | 1027 | 81.1 | 141 | 71.6 | 886 | 82.8 |  |
| 5. TV/radio |  |  |  |  |  |  | 0.511 |
| Yes | 732 | 57.8 | 118 | 59.9 | 614 | 57.4 |  |
| No | 535 | 42.2 | 79 | 40.1 | 456 | 42.6 |  |
| 6. SNS |  |  |  |  |  |  |  |
| Yes | 357 | 28.2 | 64 | 32.5 | 293 | 27.4 | 0.143 |
| No | 910 | 71.8 | 133 | 67.5 | 777 | 72.6 |  |
| 7. Parents/relatives |  |  |  |  |  |  |  |
| Yes | 480 | 37.9 | 88 | 44.7 | 392 | 36.6 | 0.033 |
| No | 787 | 62.1 | 109 | 55.3 | 678 | 63.4 |  |
| 8. Friends |  |  |  |  |  |  |  |
| Yes | 866 | 68.4 | 133 | 67.5 | 733 | 68.5 | 0.783 |
| No | 401 | 31.7 | 64 | 32.5 | 337 | 31.5 |  |
| 5) Agreement on contraceptive use by partner (n=876) |  |  |  |  |  |  |  |
| Yes | 708 | 80.8 | 162 | 82.2 | 546 | 80.4 | 0.585 |
| No | 80 | 9.1 | 19 | 9.6 | 61 | 9.0 |  |
| I don't know | 88 | 10.1 | 16 | 8.1 | 72 | 10.6 |  |

**Table 4.**
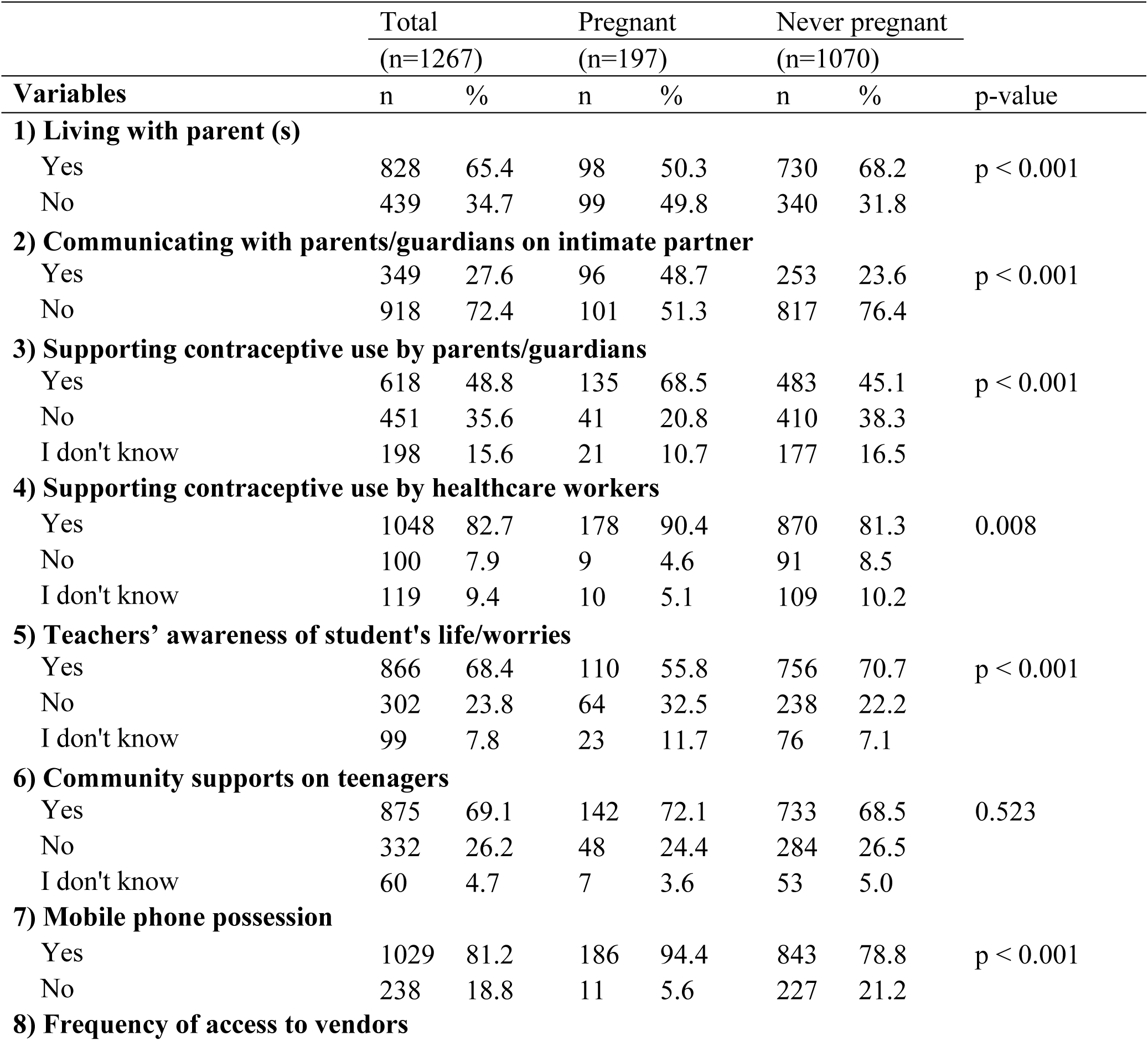
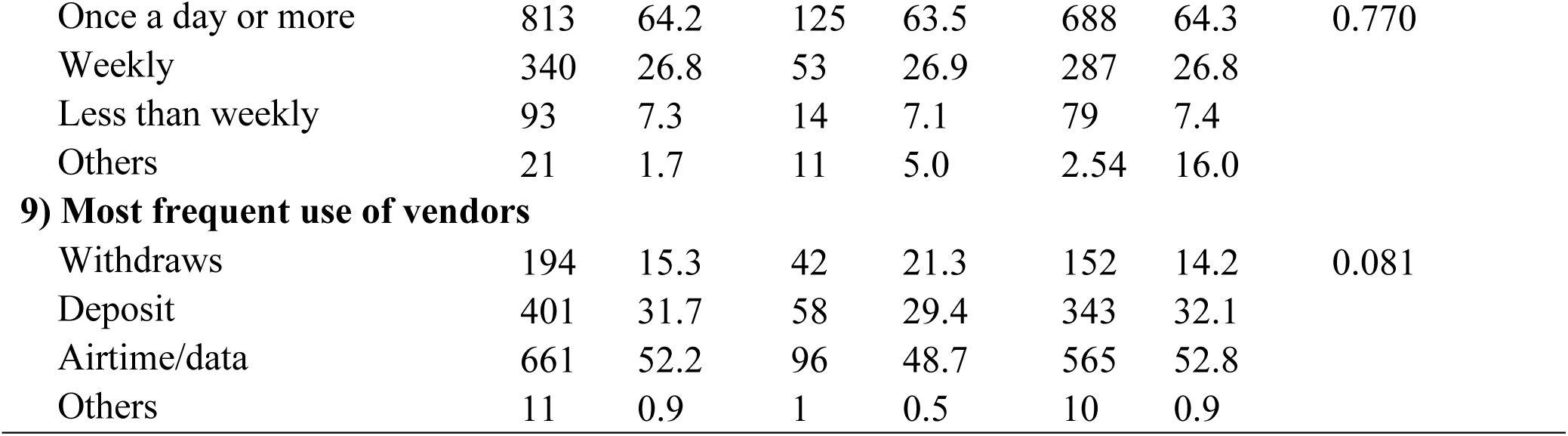
Environment related to prevent pregnancy (n = 1267)

|  | Total |  | Pregnant |  | Never pregnant |  |  |
| --- | --- | --- | --- | --- | --- | --- | --- |
|  | (n=1267) |  | (n=197) |  | (n=1070) |  |  |
| Variables | n | % | n | % | n | % | p-value |
| **1) Living with parent (s)** |  |  |  |  |  |  |  |
| Yes | 828 | 65.4 | 98 | 50.3 | 730 | 68.2 | p < 0.001 |
| No | 439 | 34.7 | 99 | 49.8 | 340 | 31.8 |  |
| **2) Communicating with parents/guardians on intimate partner** |  |  |  |  |  |  |  |
| Yes | 349 | 27.6 | 96 | 48.7 | 253 | 23.6 | p < 0.001 |
| No | 918 | 72.4 | 101 | 51.3 | 817 | 76.4 |  |
| **3) Supporting contraceptive use by parents/guardians** |  |  |  |  |  |  |  |
| Yes | 618 | 48.8 | 135 | 68.5 | 483 | 45.1 | p < 0.001 |
| No | 451 | 35.6 | 41 | 20.8 | 410 | 38.3 |  |
| I don't know | 198 | 15.6 | 21 | 10.7 | 177 | 16.5 |  |
| **4) Supporting contraceptive use by healthcare workers** |  |  |  |  |  |  |  |
| Yes | 1048 | 82.7 | 178 | 90.4 | 870 | 81.3 | 0.008 |
| No | 100 | 7.9 | 9 | 4.6 | 91 | 8.5 |  |
| I don't know | 119 | 9.4 | 10 | 5.1 | 109 | 10.2 |  |
| **5) Teachers' awareness of student's life/worries** |  |  |  |  |  |  |  |
| Yes | 866 | 68.4 | 110 | 55.8 | 756 | 70.7 | p < 0.001 |
| No | 302 | 23.8 | 64 | 32.5 | 238 | 22.2 |  |
| I don't know | 99 | 7.8 | 23 | 11.7 | 76 | 7.1 |  |
| **6) Community supports on teenagers** |  |  |  |  |  |  |  |
| Yes | 875 | 69.1 | 142 | 72.1 | 733 | 68.5 | 0.523 |
| No | 332 | 26.2 | 48 | 24.4 | 284 | 26.5 |  |
| I don't know | 60 | 4.7 | 7 | 3.6 | 53 | 5.0 |  |
| **7) Mobile phone possession** |  |  |  |  |  |  |  |
| Yes | 1029 | 81.2 | 186 | 94.4 | 843 | 78.8 | p < 0.001 |
| No | 238 | 18.8 | 11 | 5.6 | 227 | 21.2 |  |
| **8) Frequency of access to vendors** |  |  |  |  |  |  |  |
| Once a day or more | 813 | 64.2 | 125 | 63.5 | 688 | 64.3 | 0.770 |
| Weekly | 340 | 26.8 | 53 | 26.9 | 287 | 26.8 |  |
| Less than weekly | 93 | 7.3 | 14 | 7.1 | 79 | 7.4 |  |
| Others | 21 | 1.7 | 11 | 5.0 | 2.54 | 16.0 |  |
| **9) Most frequent use of vendors** |  |  |  |  |  |  |  |
| Withdraws | 194 | 15.3 | 42 | 21.3 | 152 | 14.2 | 0.081 |
| Deposit | 401 | 31.7 | 58 | 29.4 | 343 | 32.1 |  |
| Airtime/data | 661 | 52.2 | 96 | 48.7 | 565 | 52.8 |  |
| Others | 11 | 0.9 | 1 | 0.5 | 10 | 0.9 |  |

**Table 5.** Preferences for contraceptive practice (n = 1267)

|  | Total |  | Pregnant |  | Never pregnant |  |  |
| --- | --- | --- | --- | --- | --- | --- | --- |
|  | (n=1267) |  | (n=197) |  | (n=1070) |  |  |
| Variables | n | % | n | % | n | % | p-value |
| **1) Desire for knowledge of contraception** |  |  |  |  |  |  |  |
| Yes | 1177 | 92.9 | 189 | 95.9 | 988 | 92.3 | 0.192 |
| No | 53 | 4.2 | 5 | 2.5 | 48 | 4.5 |  |
| I don't know | 37 | 2.9 | 3 | 1.5 | 34 | 3.2 |  |
| **2) Desire for developing intimate relationship** |  |  |  |  |  |  |  |
| Yes | 1054 | 83.2 | 184 | 93.4 | 870 | 81.3 | p < 0.001 |
| No | 105 | 8.3 | 5 | 2.5 | 100 | 9.4 |  |
| I don't know | 108 | 8.5 | 8 | 4.1 | 100 | 9.4 |  |
| **3) Preferred source of SRHR information** |  |  |  |  |  |  |  |
| Public health facility | 505 | 39.9 | 106 | 53.8 | 399 | 37.3 | p < 0.001 |
| Others | 672 | 53.0 | 83 | 42.1 | 589 | 55.1 |  |
| I don't want to know | 90 | 7.1 | 8 | 4.1 | 82 | 7.7 |  |
| **4) Comfortable channel to obtain contraceptives** |  |  |  |  |  |  |  |
| 1. Public health facility |  |  |  |  |  |  |  |
| Yes | 952 | 75.1 | 160 | 81.2 | 792 | 74.0 | 0.032 |
| No | 315 | 24.9 | 37 | 18.8 | 278 | 26.0 |  |
| 2. Private facility |  |  |  |  |  |  |  |
| Yes | 366 | 28.9 | 58 | 29.4 | 308 | 28.8 | 0.852 |
| No | 901 | 71.1 | 139 | 70.6 | 762 | 71.2 |  |
| 3. Pharmacy/drug shop |  |  |  |  |  |  |  |
| Yes | 445 | 35.1 | 61 | 31.0 | 384 | 35.9 | 0.183 |
| No | 822 | 64.9 | 136 | 69.0 | 686 | 64.1 |  |
| 4. Community-based distributor |  |  |  |  |  |  |  |
| Yes | 53 | 4.2 | 23 | 11.7 | 30 | 2.8 | p < 0.001 |
| No | 1214 | 95.8 | 174 | 88.3 | 1,040 | 97.2 |  |
| **5) Agree on vendor as suitable channel for SRHR services (n=1264)** |  |  |  |  |  |  |  |
| Yes | 880 | 69.6 | 151 | 76.7 | 729 | 68.3 | 0.013 |
| No | 295 | 23.3 | 30 | 15.2 | 265 | 24.8 |  |
| Don't to know | 89 | 7.0 | 16 | 8.1 | 73 | 6.8 |  |

Table 6 presents factors associated with experiencing pregnancy/impregnation. Girls’ pregnancy rate was higher than the boys’ impregnation rate (adjusted odds ratio [AOR]: 2.29, 95% confidence interval [CI]: 1.52-3.44). Factors associated with the lower prevalence of pregnancy were currently student status (AOR: 0.36, 95% CI: 0.24-0.54), living with parents (AOR: 0.48, 95% CI: 0.33-0.69), and teachers’ engagement (AOR: 0.56, 95% CI: 0.37-0.84). Contrarily, factors associated with the higher pregnancy were knowledge regarding how to use contraceptives (AOR: 2.28, 95% CI: 1.16-4.49), communication with parents/guardians (AOR: 1.91, 95% CI: 1.32-2.75), supporting contraceptive use by parents (AOR: 1.64, 95% CI: 1.03-2.59), and mobile phone possession (AOR: 2.31, 95% CI: 1.09-4.90). In addition, adolescents who had experienced pregnancy were more likely to use public health facilities (AOR: 1.66, 95% CI: 1.10-2.49) and community-based distributors (AOR: 1.72, 95% CI: 1.11-2.66). Healthcare workers and community support were not correlated with the pregnancy experiences.

**Table 6.** Factors associated with experiencing pregnancy (n = 1267)

## Discussion

This study illustrates the current sexual behaviors of adolescents who used the services of vendors in Eastern Uganda. Nearly 70% of the analyzed participants had sexual intercourse. The approximate age at sexual debut was approximately 15 years for both sexes—more than 20% of those who had sexual experience had ever been pregnant. Parental engagement was a key factor associated with pregnancy. Teachers’ engagement with students’ lives reflected a reduction in pregnancy risks. In contrast, mobile phone possession and desire to get contraceptives from community-based distributors were positively correlated with pregnancy experiences.

### Active sexual behavior

Adolescents in SSA are sexually active [34, 35]. The Ugandan national average of percentage of those who had sexual intercourse among unmarried population aged 15-49 years were 59.6% for male and 45.6% of female in 2021 [9]. Our respondents showed more active sexual behaviors than the national trend, even limiting the younger population. The Ugandan national average age at first intercourse among adults aged 20-49 years was 18.3 years for males and 16.6 years for females [9]. Compared with the national average, our study participants had an early sexual initiation (15.4 years for both sexes; 5.0 years for males; 15.9 years for females). These trends were much lower than those in other SSA countries, for example, 17.0 years for females in Nigeria [27]. Furthermore, the pregnant group experienced the first sexual intercourse 0.44 years earlier than the other group in this study. This was in line with previous studies that revealed that earlier first intercourse was associated with higher unintended pregnancy rates in SSA [14, 34-36]. Our study found that the experience of pregnancy was 0.36 times lower among school students than among non-students, implying that pregnancy led to drop-out from school or out-school teens are more likely to face pregnancy risks. This supports that longer school years may reduce pregnancy risks, and extension of compulsory education years by the governmental strategy may partially effect to its purpose [14]. In addition, the Uganda government had issues several laws to reduce unintended girls pregnancy risk since 2000, such as the Unholy Law and the prohibition of marriage for girls under the age of 18 years [14]. However the data are limited to prove the effectiveness of these governmental compulsory measures [14]. Therefore, further research is needed to capture explicit adolescent needs to lower sexual activities nationwide and strengthen effective, reality-based policies [12, 14].

### Contraceptive knowledge and practice

Non-practice of contraception is a critical determinant of unintended adolescent pregnancies in SSA [8, 36]. Most respondents in this study had a basic knowledge of contraceptive methods, which is similar to the national trend among sexually active unmarried both-sex teenagers (97.8% for boys and 99.4% for girls aged 15-19 years) [26]. However, the participants’ practical knowledge on how to use the methods was 16% points lower than their basic knowledge. In addition, only 9.1% of the adolescents had correct knowledge of the fertile period of the menstrual cycle, which was lower than the national average among girls aged 15-19 years (13.8%) in 2016 [26]. Our descriptive analysis revealed that adolescents with pregnancy experience tended to have more knowledge and access to contraception and support from parents/guardians and healthcare workers. Although this cross-sectional study could not identity causality, the results may be interpreted that teens those who experienced pregnancy became more serious about searching for information and mobilizing services after pregnancy. It can be said that Uganda’s SRHR education either at school, community, nor home does not provide preventative information of pregnancy and meet their real needs, although the Ministry of Education and Sports encourages the improvement of SRHR education in schools. In our study, the most common source of contraceptive information was schools; however, this was not the preferred source of SRHR information. A previous study concluded that insisting on abstinence in school education was ineffective in preventing teenage pregnancies [37]. Providing practical knowledge on sex, pregnancy, contraception, and sexually transmitted infections is crucial among adolescents at the scale, through home, school, and community [37-39]. Some studies emphasized that younger people were exposed to more unsafe sexual intercourse and were flexible in absorbing concepts related to gender, suggesting that sex education from an early age before sexual initiation was effective [36, 40-42]. A study on children aged 10-14 years in Uganda supported this suggestion [43]. Education content should include sex, pregnancy, delivery, sexually transmitted infections, other health risks, the efficacy of vulnerable populations, and future income potential [3, 7, 11, 32, 37, 38, 44]. The content should be designed according to the needs of different adolescent age groups [41, 45]. About half of those who had ever been pregnant were most likely to terminate their pregnancies. Since abortion for non-medical reasons is illegal in Uganda, statistics on abortion data were collected only through post-abortion care at health facilities and the actual situation is underreported. The previous qualitative studies captured that many girls have used traditional medicines or unsafe means to terminate unwanted pregnancies and received no after-abortion care [11, 12, 14]. Ignoring high-risk, unsafe self-abortion or necessary after-abortion care is likely to lead to maternal death and adverse maternal outcomes [12]. Sexual education emphasizing the risks of abortion and infertility is needed [11, 37].

### Parental involvement

Parental involvement was an essential factor influencing adolescent unintended pregnancy in several studies in SSA countries [31, 32, 38]. In this study, "living with parents" was indicated to be a deterrent factor to sexual behavior. Studies in Ghana have reported a similar situation in which parental involvement reduced pregnancy occurrence [31, 38]. Conversely, parental interaction and parental contraceptive support were positively correlated with pregnancy experience, which may imply that parents’ substantive care and support increased after their children’s pregnancy experience, similar to the results of a Ghanaian study [31]. Parents’ neglect of their children and hesitancy to teach sex education because they did not know how to interact with them was also noted in a previous study in Uganda [11]. Parents’ motivation for sex education for their children, their own acquisition of correct SRHR knowledge, and their ability to interact with the children need to be improved [27, 32, 38]. Urgent development of educational program guidance for families (parents/guardians) and communities, including practical sex education content and skills to teach children in Uganda [11] is required.

### Contraceptive accessibility in the community

The prevalence of unintended pregnancies is closely related to access to contraceptives [44]. Paid services at private clinics or pharmacies were hurdles for teenagers in Uganda and they tend to avoid public health facilities because they follow formal procedures, ask detailed questions about personal information and the purpose of contraceptive use, and, in some cases, may be known to their parents [11, 12]. This study found that motivation for contraceptive use was higher among those who had experienced pregnancy, and in fact, they mobilized a variety of contraceptive sources. These facts imply that teenagers who experienced pregnancies have strengthened their contraceptive capabilities. A Ghanaian study also pointed out that many girls who had experienced pregnancy developed competencies to deal with pregnancy prevention [31]. Our findings also indicated that easily accessible community-based distributors showed a preference for and a higher desire for future use among teenagers who had experienced pregnancy.

This study also found Approximately 70% of adolescents thought that the vendor was a suitable channel for receiving SRHR services. Furthermore, adolescents who have experienced pregnancy are more likely to prefer accessing vendor-based SRHR service. These facts suggest that vendors may be effective channels for providing SRHR services, especially for high risk population. Chandra-Mouli pointed out that interventions implemented in many programs such as facility-based interventions, including school education, health center activities, and youth centers with peer-educators in the community, have not been cost-effective or sustainable [19]. The Ugandan government has called for innovative approaches that utilize local mechanisms outside the health and education sectors [46, 47]. In response to the global community calling for innovative platforms to reach vulnerable populations nearly a decade ago [4, 17, 21], our results suggest that vendors in Uganda could be a cost-effective platform to reach adolescents and strengthen their competencies in accessing contraceptives and practice.

## Limitation

This study has several limitations. First, we collected data from adolescents who used the vendor services, meaning that our findings could not represent the general population. Second, since the survey was dependent on self-reporting, the study is susceptible to recall and social desirability biases. Third, the nature of the quantitative study and cross-sectional design could not describe causality of pregnancy, and limited further interpretation of underlying social, cultural, and historical factors. Fourth, our study did not explore the economic status of the adolescents because of difficulties in accessing accurate data and comparing adolescents living with their parents and others using a structured questionnaire. In addition, we did not include patients with human immunodeficiency virus (HIV) acquired immunodeficiency syndrome (AIDS) in this study. As there is still high stigma attached to HIV/AIDS status in Uganda, we determined that obtaining such information through self-reports would not provide reliable information. To overcome these limitations, a qualitative study with a diverse population of children, parents, and social decision-makers is required. Despite these limitations, to the best of our knowledge, this is the first community-based quantitative study of sexual behaviors and contraception preferences among unmarried girls and boys aged 15-19 years in Uganda and identified the unmet needs of sexual and reproductive health services.

## Conclusions

This study revealed a picture of early sexual debut and pregnancy risk among adolescents aged 15-19 years in Eastern Uganda. This study found that the important factors underlying such situations were a lack of knowledge of pregnancy risk and correct knowledge of contraception as well as a lack of practical sexual education by parents and schools. However, there was light on the existence of a contemporary community resource with a high affinity for adolescents, known as mobile money vendors. Young vendors who are in the community, geographically accessible, and have a close relationship with adolescents could be a potential channel for delivering necessary information on pregnancy risk and promoting safe contraceptive use among adolescents. Further studies are needed to develop a model for promoting modern contraceptive use among vendors, test its scalability, and share the results with policymakers in Uganda.

## Data Availability

All data produced in the present study are available upon reasonable request to the authors.

https://www.umin.ac.jp/icds/index.html

## List of abbreviations

AIDS: acquired immunodeficiency syndrome
AOR: adjusted odds ratio
CI: confidence interval
HIV: human immunodeficiency virus
SRHR: sexual and reproductive health and rights
SSA: Sub-Saharan Africa

## Declarations

### Ethics approval and consent to participate

Ethical approval was obtained from the Ethics Review Committee of Uganda Christian University (UCUREC-2023-710), Uganda National Council for Science and Technology (SS2284ES), and Ethics Review Committee of the JICA Ogata Sadako Research Institute for Peace and Development in Japan (JICA Ogata RI, reference: JICA [DI] 202312220002). Written informed consent was obtained from all adolescent participants and the parents/guardians of adolescents aged 15-17 years.

### Consent for publication

Not applicable.

### Availability of data and materials

The dataset supporting the conclusions of this article is available in the repository of the University Hospital Medical Information Network (UMIN000053332) dataset in https://rctportal.niph.go.jp/en.

### Authors’ contributions

MK, MS, RS, KS, SG and AMN were involved in the development of the conception and study design. MK, MS, and RS were responsible for field research implementation. SH provided technical advice on the overall methodology, including the sampling. MK was involved in drafting the manuscript. PW supervised the study and was involved in critical revision of the manuscript. MS, RS, MKM and AMN provided technical comments on the manuscript draft. MK was responsible for the fund. All authors were involved in the final approval of the manuscript and the decision to submit the manuscript for publication.

## Acknowledgements

We would like to express our gratitude to all study participants, the Ministry of Health for their invaluable support. Our heartfelt thanks also go to all members of the research team of the Busoga Health Forum, whose efforts facilitated the implementation of this study, including the development of a network involving various stakeholders and healthcare facilities.

## Authors’ information

^1^Ogata Sadako Research Institute for Peace and Development, Japan International Cooperation Agency, Tokyo, Japan. ^2^Department of Global Health Research, Juntendo University, Tokyo, Japan. ^3^School of Tropical Medicine and Global Health, Nagasaki University, Nagasaki, Japan. ^4^Graduate School of Biomedical Sciences, Nagasaki University, Nagasaki, Japan. ^5^School of Biomedical Sciences, Makerere University, Kampala, Uganda. ^6^School of Public Health, Makerere University, Kampala, Uganda. ^7^Iganga Health District office, Iganga, Uganda. ^8^Busoga Health Forum, Jinja, Uganda.

